# COVID-19 infection during pregnancy: a systematic review to summarize possible symptoms, treatments, and pregnancy outcomes

**DOI:** 10.1101/2020.03.31.20049304

**Authors:** Md. Mostaured Ali Khan, Md. Nuruzzaman Khan, Md. Golam Mustagir, Juwel Rana, Md. Rajwanul Haque, Md. Mosfequr Rahman

**Affiliations:** Department of Population Science and Human Resource Development, University of Rajshahi, Rajshahi-6205, Bangladesh; School of Public Health and Medicine, Faculty of Health and Medicine, The University of Newcastle, Australia; Department of Population Sciences, Jatiya Kabi Kazi Nazrul Islam University, Mymensingh, Bangladesh; Department of Public Health, School of Health and Life Sciences, North South University, Bashundhara, Dhaka-1229, Bangladesh; Department of Biostatistics and Epidemiology, School of Public Health and Health Sciences, University of Massachusetts Amherst, Amherst, MA, United States; MEL and Research, Practical Action, House no. 28/A, Road no. 5, Dhanmondi, Dhaka-1205, Bangladesh

**Keywords:** Coronavirus, 2019-nCoV, COVID-19, Pregnant women, pregnancy

## Abstract

**Background:** With the exponential increase in coronavirus disease 2019 (COVID-19) worldwide, an increasing proportion of pregnant women are now infected during their pregnancy. The aims of this systematic review were to summarize the possible symptoms, treatments, and pregnancy outcomes of women infected with COVID-19 during their pregnancy.

**Methods:** Four databases (Medline, Web of Science, Scopus, and CINAHL) were searched on March 25, 2020, using the following keywords: “COVID-19”, “nCoV-2019”, and “coronavirus.” Articles included if they reported either the symptoms, treatments for the women who had been infected with the COVID-19 during their pregnancy or pregnancy outcomes. The selected articles’ results were summarized employing a narrative synthesis approach.

**Results:** A total of nine studies were selected for this study, comprising 101 infected pregnant women. Other than the infected general population, infected pregnant women reported different symptoms; however, fever (66.7%), cough (39.4%), fatigue (15.2%), and breathing difficulties (14.1%) were common. Infected pregnant women were given different treatments than the general infected population. The C-section was a common (83.9%) mode of delivery among infected pregnant women, and a higher proportion of births were preterm births (30.4%) and low birth weight (17.9%).

**Conclusions:** Pregnant infected women had different symptoms, and they were given dissimilar treatments than the general infected population. Healthcare providers may have appropriately informed about these symptoms and treatments. They, therefore, would be able to handle infection during pregnancy effectively, which would reduce common adverse consequences among infected pregnant women.

## Introduction

The world is now facing a new coronavirus disease 2019 (COVID-19, started in December 2019 in Wuhan, China), a major epidemic threat that the world has ever faced [1]. Since then, the virus has spread to 205 countries or territories and infected around 1.2 million people, and around 65 thousand of them died as of April 5, 2020 [2]. These rates of infections and deaths have been changing daily because of their higher transmission capacity. Consequently, the witness countries’ healthcare systems are now facing problems to treat the infected people properly, although many of them are critically infected and need comprehensive care (5%) [3, 4]. Essentially, these increasing rates are reported when the World Health Organization (WHO) has declared COVID-19 as a global pandemic and asked countries to take aggressive measures that are even reflected in countries’ various initiatives [5, 6]. The roots of transmission of this virus from an infected to non-infected person are in contact with an infected person’s respiratory droplets (coughs or sneezes) [7], and touching surfaces or objects that were touched by the infected person [7, 8], and has the capacity to infect 2.28 persons per day [9].

The virus is prevalent in all age clusters, irrespective of sex. However, it could cause more adverse consequences, including deaths for persons who have pre-existing non-communicable diseases, including diabetes, hypertension, chronic respiratory diseases, and cancer [1]. Notably, the majority of deaths reported worldwide have occurred among people who had a history of one or more morbidities, which are often common among the elderly [1, 10]. The causes of death are many, but poor immunity capacity of people with one or more morbidities might be the main.

Women during pregnancy often face several pregnancy-related complications and more susceptible to respiratory pathogens that may put them at a higher risk of adverse consequences. Getting COVID-19 infected at this stage, therefore, may put them in further risk of occurring adverse pregnancy and newborn outcomes, including deaths; however, the estimate is lacking [11]. Similar to the current COVID-19, severe acute respiratory syndrome (SARS) was reported in 2007 and showed devastating consequences if it occurred during pregnancy [9, 12]. Earlier diagnosis of COVID-19 infection status, a clear understanding of all possible symptoms of infection during pregnancy, and proper treatment are important to combat the adverse consequences.

The available studies on the symptoms of COVID-19 were mainly conducted in the general infected people, and fever, cough, fatigue, and shortness of breath were the common symptoms among them [13-18]. These symptoms reported in a few relevant studies vary for pregnant women, which are inconsistent across studies. For instance, Liu et al. [19] identified cough, shortening of breath, and fatigue alongside fever as the most important symptoms of COVID-19 among pregnant women, whereas cough and fever were reported as vital symptoms of COVID-19 in the Zhu et al. [20] study. The treatments provided to the COVID-19 infected pregnant women were also varied. Oxygen support, antiviral therapy, and antibiotic therapy were the treatments provided to the infected pregnant women mentioned in Liu et al. [21] study. Alternatively, Zhu et al.’s study [20] reported that antibiotic therapy was avoided for all infected pregnant women, but oxygen support and antiviral therapy were provided. Under these circumstances, summarizing all possible symptoms and treatments are necessary to assist healthcare personnel. They, thus, would able to make evidence-based decisions to identify infected pregnant women early and provide the most effective treatments. Moreover, knowledge of the possible adverse outcomes would help healthcare personnel to take precautionary measures earlier. Therefore, this study summarized the symptoms, treatments, and pregnancy outcomes of women who were infected with COVID-19 during their pregnancy.

## Methods

A systematic review following the Preferred Reporting Items for Systematic Review and Meta-analysis (PRISMA) consensus statement was conducted [22]. Relevant and available studies related to COVID-19 infection among pregnant women were included.

### Search strategy

Systematic computerized literature searches of the Medline, Web of Science, Scopus, and CINAHL databases were conducted on March 25, 2020. Studies published since the COVID-19 started were included. Searches were conducted based on individual comprehensive search strategies for each database. We developed a search strategy consisting of free-text words, words in title/abstract, and medical subject headings (MeSH), combined using Boolean operators (AND, OR). We also searched the reference list of the included articles and selected journal websites.

### Study selection criteria

Studies were selected based on the inclusion and exclusion criteria outlined below.

#### Inclusion criteria

Studies included if they reported COVID-19 infection among pregnant women and the following: symptoms of the infection, treatment given, and outcome of pregnancy (if the delivery had occurred). Studies published in the English language worldwide, irrespective of study design, were included.

#### Exclusion criteria

Studies excluded if they reported COVID-19 infection among non-pregnant women or general patients and wrote in languages other than English. We also excluded review papers and studies in which the exposure status (COVID-19 among pregnant women), symptoms, treatments, and outcomes were not clearly reported.

### Data extraction and analysis

Two authors (MMAK and MGM) extracted information using a pre-designed, trailed, and modified data extraction sheet. The extracted information included year of publication, study location, study design, study sample size, symptoms reported, treatment given, and pregnancy outcome (if delivery had occurred). The corresponding author solved any disagreements on information extraction. The modified Newcastle-Ottawa scale, as part of the data extraction strategy, was used to assess study quality.

The information recorded was dichotomous in nature. We, therefore, used a narrative synthesis approach to summarize the findings from all the retrieved studies. All symptoms following COVID-19 infection among pregnant women, the treatments given to them, and the outcomes of their pregnancy are presented in detail along with relevant data if available.

## Results

### Study selection

A total of 1,818 articles dating from the inception of the COVID-19 were identified, of which 1,706 articles were excluded based on the title, abstract, and relevance of the research questions. One hundred twelve articles were selected for full-text review, of which 103 articles were excluded because study samples were general infected population (95), reported different outcomes (4), and written in languages other than English (2). A total of 9 articles were finally selected for this study.

The background characteristics of the selected studies are summarized in Table 1. All included studies were conducted in China following the initiation of the COVID-19 outbreak in December 2019. Five of the nine included studies were followed retrospective cohort study design, two were cross-sectional, and two were a case report. A total of 101 women were included in the selected nine articles; their mean age was 30 years, and most of them were in the third trimester of pregnancy (gestational age 22-41 weeks).

**Table 1.** Background characteristics of the selected studies.

| SL no. | Authors | Study design | Study location | Study date | Study sample size | Mean age of study sample, (±SD) | Gestational weeks at admission (in weeks) |
| --- | --- | --- | --- | --- | --- | --- | --- |
| 1 | Chen et al. [43] | Retrospective cohort | Wuhan, China | 12 Feb, 2020 | 9 | 29.9 (±2.9) | 36-40 |
| 2. | Liu et al. [21] | Cross sectional | Hubei, China | 7 Mar, 2020 | 15 | 32.0 (±5.0) | 12-38 |
| 3. | Liu et al. [19] | Cross sectional | Hubei, China | 11 Mar, 2020 | 41 | NA | 22-40 |
| 4. | Liu et al. [44] | Retrospective cohort | Hubei, China | 28 Feb, 2020 | 13 | 29.7 (±2.1) | 25-38 |
| 5. | Wang et al. [45] | Case report | Suzhou, China | 28 Feb, 2020 | 1 | 28 (±NA) | 30 |
| 6. | Chen et al. [46] | Retrospective cohort | Hubei, China | 16 Mar, 2020 | 4 | 29 (±3.9) | 37-38 |
| 7. | Zhu et al. [20] | Retrospective cohort | Hubei, China | 10 Feb 2020 | 10 | 30.7 (±1.8) | NA |
| 8. | Yu et al. [47] | Retrospective cohort | Wuhan, China | 24 Mar, 2020 | 7 | 32.1 (±1.5) | 37-41+ |
| 9. | Zambrano et al. [48] | Case report | Honduras | 25 Mar, 2020 | 1 | 41 | 31 |

**Note:** NA: Not Available

### Symptoms of COVID-19 among pregnant women

Table 2 presents the symptoms reported in the included studies. Where available, the symptoms were reported on case-basis included in the studies. Fever (66.7%) and cough (39.4%) were the major symptoms following COVID-19 infection among pregnant women **(Figure 2)**. The other symptoms reported were fatigue (15.2%), breathing difficulties (14.1%), and myalgia (7.1%). Sore throat (5.1%), diarrhea (4%), poor appetite (3%), headache (3%), and malaise (2%) were also reported among pregnant women following COVID-19 infection. Two different pregnant women also reported mulligrubs and cholecystitis, respectively, which were added to the common symptoms.

**Table 2.**
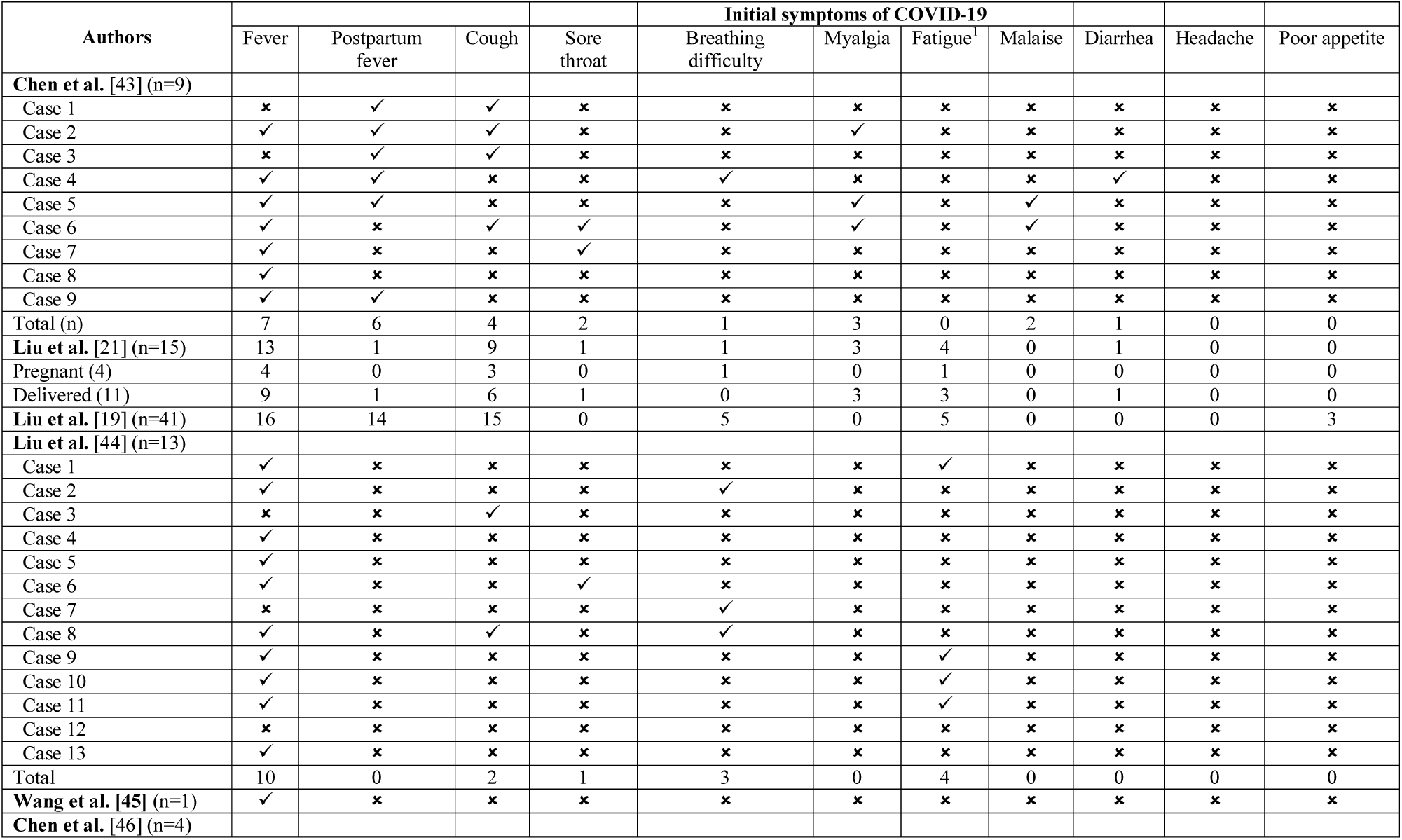
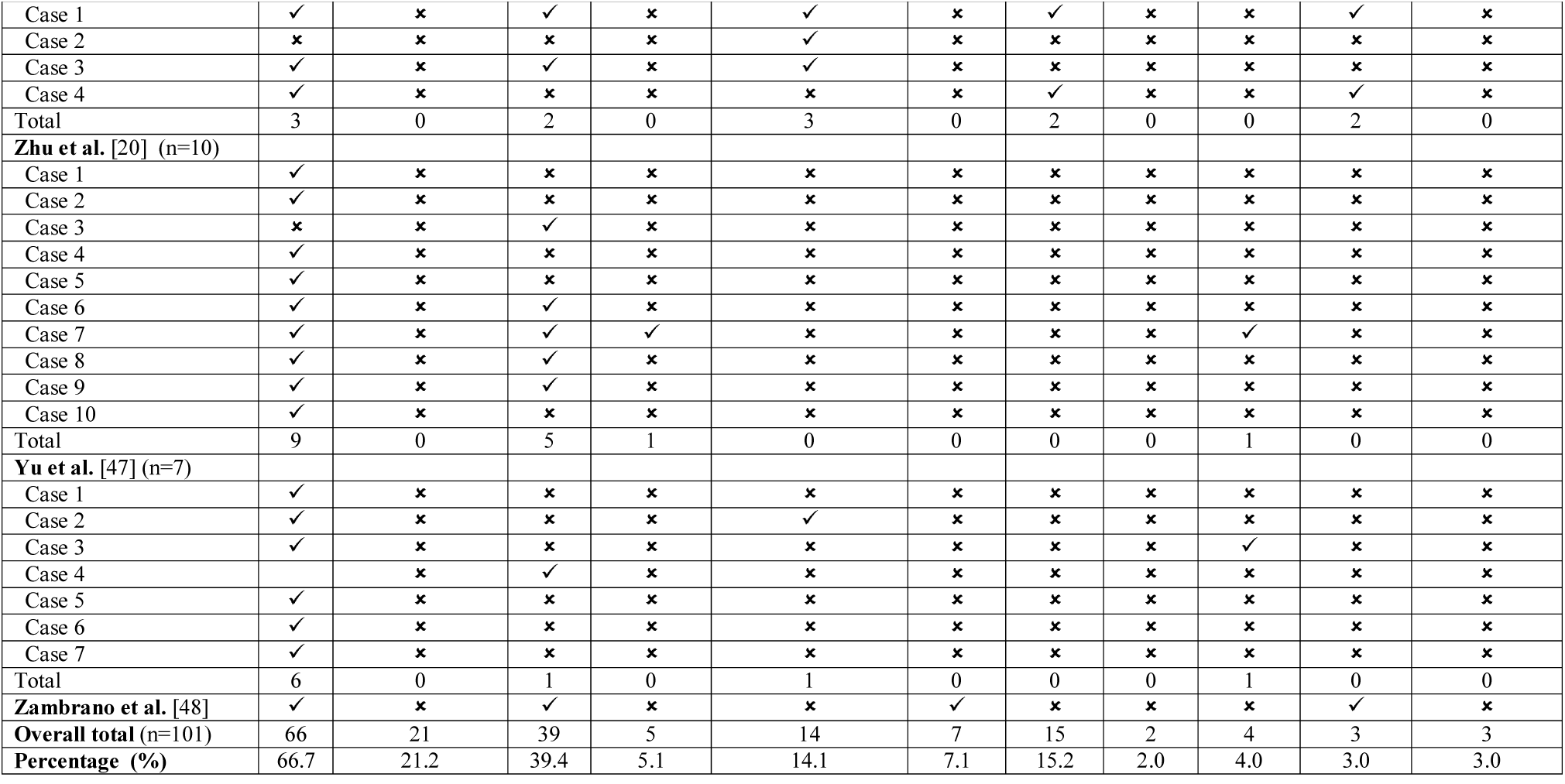
Detailed case specific symptoms of the COVID-19 infection among pregnant women.

**Figure 1.**
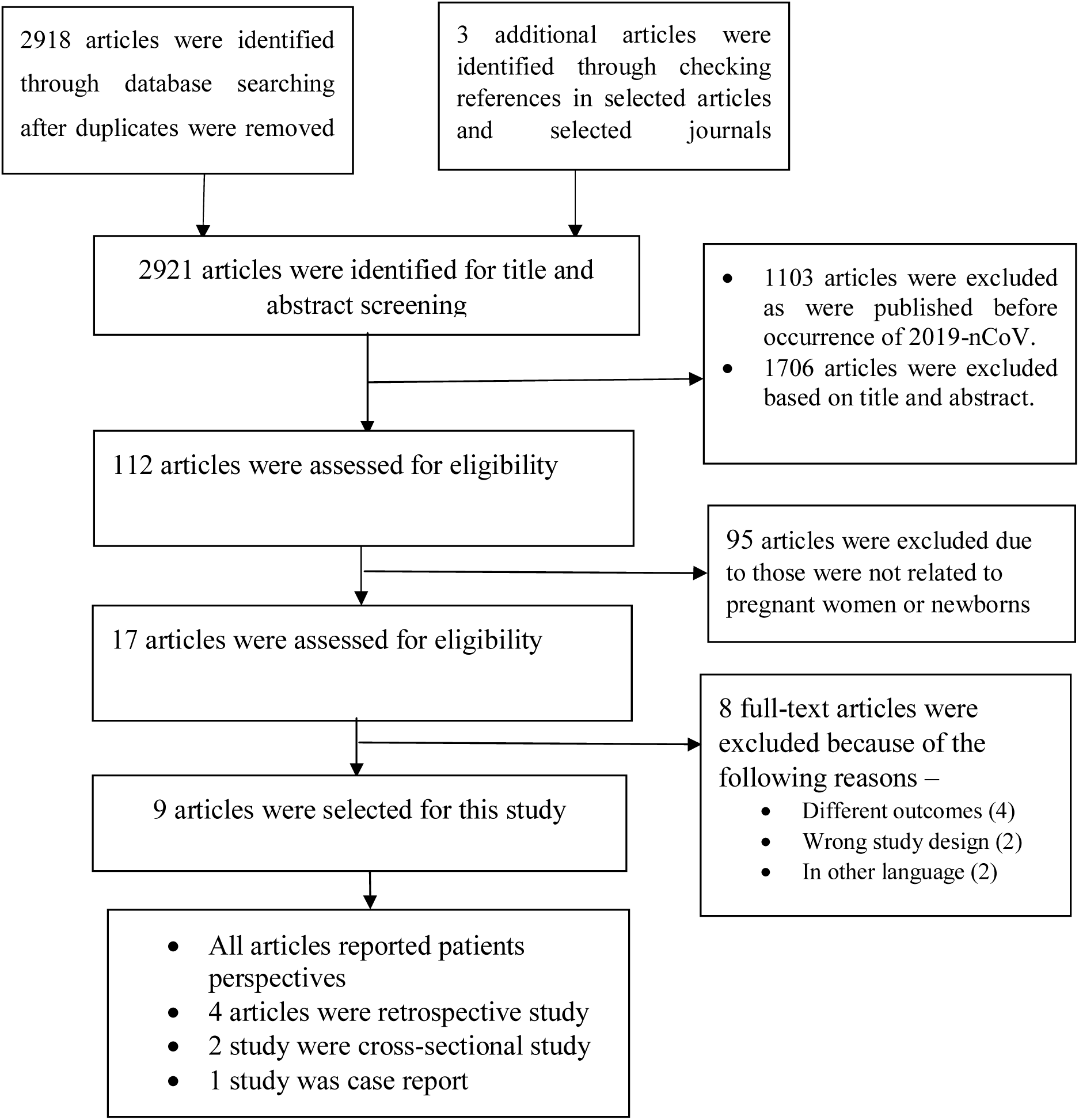
Schematic representation of studies included in the systematic review using PRISMA checklist and flow diagram.

**Figure 2.**
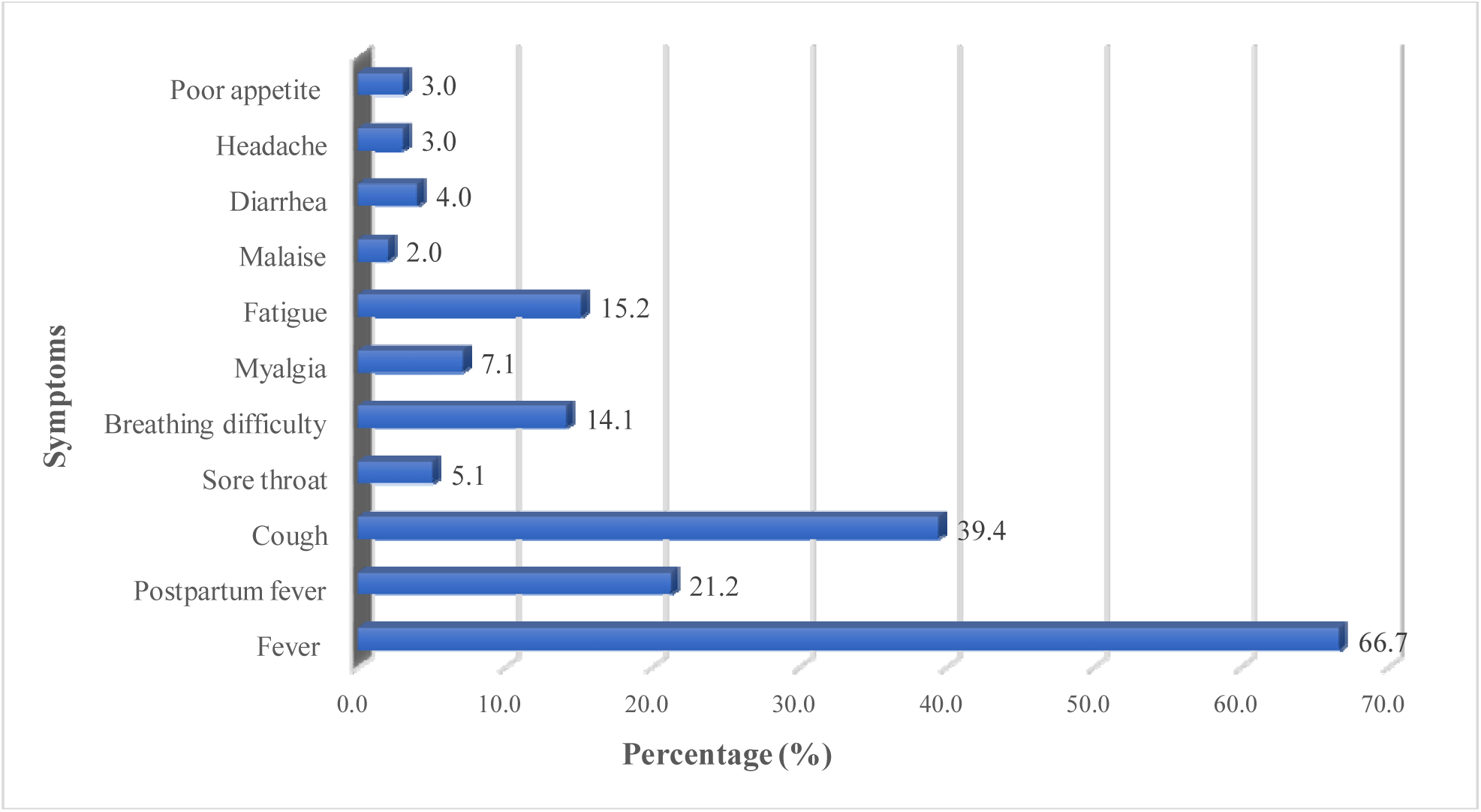
Symptoms of the COVID-19 infection during pregnancy.

### Treatments used to treat COVID-19 diseases among pregnant women

The treatment patterns of COVID-19 infection among pregnant women during their pregnancy or following delivery are summarized in Table 3. Of the nine articles included, four articles reported similar treatments of oxygen and antibiotic therapy. Antiviral therapy was also used in some cases. Cefoperazone sodium, cephalosporins, quinolones, and macrolides were the commonly used antibiotics in antibiotic therapy, and lopinavir (400mg), ritonavir (100mg), arbidol, and oseltamavir were commonly used in antiviral therapy.

**Table 3.** Treatments given to pregnant women following the COVID-19 infection and the delivery methods (if pregnancy had been ended with delivery).

| Studies | Overall treatment |  |  |  |
| --- | --- | --- | --- | --- |
|  | Oxygen support | Antiviral therapy<sup>1</sup> | Antibiotic therapy<sup>2</sup> | Corticosteroid |
| **Chen et al. [43] (n=9)** |  |  |  |  |
| Case 1 | ✓ | ✓ | ✓ | ✗ |
| Case 2 | ✓ | ✓ | ✓ | ✗ |
| Case 3 | ✓ | ✓ | ✓ | ✗ |
| Case 4 | ✓ | ✗ | ✓ | ✗ |
| Case 5 | ✓ | ✗ | ✓ | ✗ |
| Case 6 | ✓ | ✗ | ✓ | ✗ |
| Case 7 | ✓ | ✓ | ✓ | ✗ |
| Case 8 | ✓ | ✓ | ✓ | ✗ |
| Case 9 | ✓ | ✓ | ✓ | ✗ |
| **Total (n)** | 9 | 6 | 9 | 0 |
| **Liu et al. [21] (n=15)** |  |  |  |  |
| **Pregnant (n=4)** | 3 | 0 | 4 | 0 |
| **Delivered (n=11)** | 11 | 11 | 11 | 0 |
| **Liu et al. [19] (n=41)** | - | - | - | - |
| **Liu et al. [44] (n=13)** | - | - | - | - |
| **Wang et al. [45] (n=1)<sup>a</sup>** | ✓ | ✓ | ✓ | ✗ |
| **Chen et al. [46] (n=4)** | - | - | - | - |
| **Zhu et al. [20] (n=10)** | 2 | 5 | 0 | 0 |
| **Yu et al. [47] (n=7)<sup>b,c</sup>** | ✓ | ✓ | ✓ | ✗ |
| **Zambrano et al. [48]** | - | - | - | - |

**Note:** <sup>1</sup>Specific treatment: Arbidol, Oseltamivir, Lopinavir, Ritonavir, ganciclovir, interferon was given in the study of Wang et al. [45], Zhu et al. [20], and Yu et al. [47]; <sup>2</sup>Specific treatment: Cefoperazone Sodium, Cephalosporins, Quinolones, Macrolides, Sulbactam Sodium (anti-bacterial treatment) (Wang et al. and Zhu et al.); <sup>a</sup>Human Serum Albumin based delivery; <sup>b</sup>Traditional Chinese medicine was also used; <sup>c</sup>two patients were treated with a single antibiotic and five patients were given combination therapy

### Pregnancy outcomes following delivery among COVID-19 infected women

The maternal and newborn outcomes following COVID-19 infection during pregnancy are summarized in Table 4. This study included 101 women, of which delivery-related information was available for 60 women; 56 had given live birth, and the remaining four were still pregnant at the time of study conducted. Of all deliveries occurred, 83.9% had gone through the C-section, and around 30.4% of the total deliveries were premature. Among these reviewed cases, one maternal death and one neonatal death were also reported following COVID-19 infection. The birth weight of the babies was normal in most cases, although 17.9% of the newborns had low birth weight (LBW). Only one of the included studies reported evidence of mother-to-child transmission of COVID-19 infection after 36 hours of delivery.

**Table 4.**
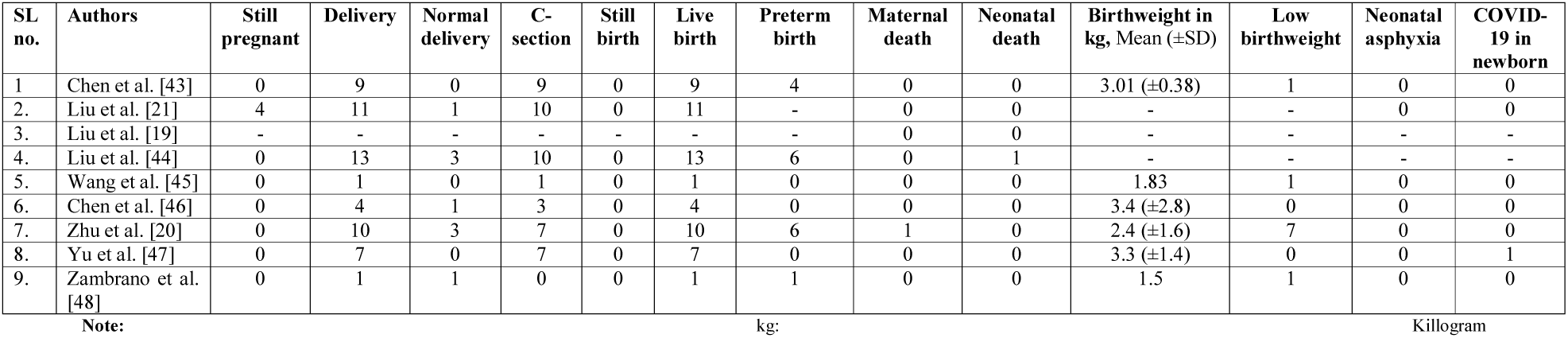
Maternal and newborn adverse outcomes following the COVID-19 infection among pregnant women

## Discussion

The world is now experiencing an exponential increase in the COVID-19 infected people, and a significant proportion of them are pregnant women. We summarized the symptoms following COVID-19 infection among pregnant women, and the treatments are commonly provided to them and the outcomes of the pregnancy. A total of nine studies were included in this review, in which 101 infected pregnant women data were analyzed, and the majority of them were in the third trimester of pregnancy. All included studies were conducted in China. Symptoms reported by pregnant women following COVID-19 infection were slightly different from the common symptoms in general infected people, but fever and cough were common. Oxygen and antibiotic therapy were given as treatments for most infected pregnant women. Of the sample analyzed in the eight studies included, 56 of them gave babies following the infection occurred. Approximately 84% of the infected pregnant women had gone through the C-section; around 30% of women had given premature birth, and a total of 18% of these babies were born with LBW. These summary findings will assist healthcare personnel for the better management of pregnant women who have been infected with COVID-19, which potentially reduces adverse consequences for women as well as their babies.

Several questions have been provoked regarding maternal and neonatal safety if women were infected with COVID-19 during pregnancy. The causes are complications following getting infected and evidence of higher adverse consequences if it has happened with existing morbidity, and these could be added to the usual pregnancy and delivery-related complications. These factors may increase the occurrence of adverse maternal health and birth outcomes, although estimates are lacking. Notably, earlier detection of the infection through tests, symptoms, and effective treatments could reduce these risks. As revealed in this study, based on available published research, fever, cough, fatigue, and breathing difficulties are common symptoms among both pregnant women and infected general people following COVID-19 infection [23, 24]. Although most of the symptoms were almost similar, infected pregnant women had some additional symptoms such as sore throat, myalgia, and poor appetite. Postpartum fever was also highly observed in women following delivery. Therefore, close monitoring of these symptoms could be an effective way of early detection of COVID-19 infection during pregnancy. Previous corona-like virus outbreaks, such as infection of SARS during pregnancy, reported malaise, chills, and rigors as common symptoms along with fever, cough, and breathing difficulties [25].

Given that no specific treatments or vaccines have been discovered yet, the COVID-19 infection during pregnancy would put healthcare providers in additional challenges [23]. This is because of the possible adverse effects of the medicine used on the fetus as well as pregnancy. For instance, there is evidence that medicines such as chloroquine and hydroxychloroquine are currently recommended for patients with COVID-19 [26], which may cause fetal harm [27] and adversely affect newborns through transferring from mother body by breastmilk [28, 29]. Therefore, cautions are needed in the treatment of infected pregnant women. This study revealed that most infected pregnant women were given symptomatic and supportive treatments considering pregnancy, although anti-inflammatory and antiviral treatments have been used in some cases [30, 31]. Significantly, this study summarized the commonly given antiviral treatments to the infected pregnant women were oseltamivir, lopinavir (400mg), and ritonavir (100mg), which are different from usually recommended medicines to treat COVID-19 in the general population [26]. Therefore, healthcare providers may have to be careful about given antiviral treatments to infected pregnant women.

Additionally, oxygen therapy and antibiotic therapy (e.g., cefoperazone sodium) have been used in some cases [31, 32]. Notably, further cautious measures are needed to choose the medicine from previous experience with SARS and its treatments, which is considered like COVID-19 infection, and there are many similarities in treatments given for these infections. For instance, oxygen and antiviral therapy (drugs, including ribavirin, ritonavir, and lopinavir) are used in both SARS and COVID-19 [33]. However, corticosteroids were not used to treat COVID-19, although it was used to treat SARS among pregnant women. There is evidence that the use of corticosteroids during pregnancy increases the risk of preterm birth, low birth weight, and preeclampsia [34].

Additional important findings of this study were the higher occurrence of adverse maternal newborn outcomes among COVID-19 infected mothers. For example, as this study reported, around 84% of the infected mothers had birth reported use of the C-section; a rate that is significantly higher than the usual WHO’s recommendation of 1-5% use of C-section to avoid death and severe morbidity in mothers and newborns [35, 36]. Getting COVID-19 infection increases complex viral infection among women in pregnancy; therefore, C-section is recommended to reduce perinatal and neonatal adverse outcomes [37]. We also confirm the evidence of the higher occurrence of preterm birth and LBW among the COVID-19 infected mothers than their counterparts. These associations are unique, as the previous round of viral infection, such as SARS, did not find any evidence of the association between infection during pregnancy and occurrence of LBW and preterm birth [38-41]. Viral infection affects pregnancy and fetal growth by gaining access to the placenta and decidua via hematogenous transmission or the lower reproductive tract, leading to the occurrence of such adverse outcomes [42]. However, effects can vary across cell types, gestational age, and changes in the uterine environment and maternal immunity [42]. Therefore, earlier preparedness of the healthcare sectors to handle these adverse consequences would be helpful to reduce further adverse outcomes, including maternal and perinatal mortality.

This study has several strengths and limitations. This is the first study of its kind that highlight the symptoms, treatments, and pregnancy outcomes among women who have been infected with COVID-19. These findings may help healthcare providers to take proper initiatives. We did not set any time and study design restrictions that allowed us to include a higher number of studies. However, the major limitation is generalizability because all included studies were conducted in China. Different treatments may be used to treat COVID-19 during pregnancy in other countries. Moreover, none of the included studies reported quantitative data that restrict us to conduct a narrative synthesis of the selected study’s findings rather than given any pool estimates. Despite these limitations, this study has enough merits, which will make healthcare providers informed about the symptoms, treatments, and possible outcomes in pregnant women detected with COVID-19.

## Conclusion

This study confirms that fever, cough, and breathing difficulties are the major symptoms of COVID-19 infection among pregnant women, which are similar to general infected patients. However, some additional symptoms among pregnant women are postpartum fever and breathing difficulties. Recommended treatments such as chloroquine and hydroxychloroquine for the infected people are not applicable for pregnant women because of their potential adverse effects on the fetus and newborn; therefore, supportive and symptomatic treatments are given to them. COVID-19 infection during pregnancy also increases the risk of several adverse outcomes, including higher rates of cesarean delivery, low birth weight, and preterm birth. Healthcare providers may consider these for effective management of COVID-19 infected pregnant women, which would reduce pregnancy-related adverse consequences, including maternal and newborn morbidity and mortality.

## Data Availability

All data are available in the manuscript

## Acknowledgments

The authors are grateful to the authors of the paper included in this review.

## Authors Contribution

Conceptualization: MMAK

Research design: MMAK and MNK

Data curation: MMAK and MGM

Analysis: MMAK, MGM, and MNK

Draft preparation: MMAK, MNK, MGM, JR, MMR, and MRH

Supervision: MNK

Critical review and edits: JR, MNK, MMR, and MRH

## References

1. WHO. Coronaviruses (COVID-19). 2020 [cited 2020 25.03.2020]; Available from: https://www.who.int/emergencies/diseases/novel-coronavirus-2019

2. WHO, Novel Coronavirus (COVID-19) Situation. 2020, World Health Organization, Geneva, Switzerland.

3. Fisher D and Heymann D, Q&A: The novel coronavirus outbreak causing COVID-19. BMC Medicine, 2020. 18(1): p. 57.

4. Worldmeter. COVID-19 Coronavirus Pandemic (Real time database). 2020 03.04.2020]; Available from: https://www.worldometers.info/coronavirus/.

5. Gilbert M, et al., Preparedness and vulnerability of African countries against importations of COVID-19: a modelling study. The Lancet, 2020. 395(10227): p. 871–877.

6. Bedford J, et al., COVID-19: towards controlling of a pandemic. The Lancet, 2020.

7. CDC. Coronavirus Disease 2019 (COVID-19). 2020 [cited 2020 20.03.2020]; Available from: https://www.cdc.gov/coronavirus/2019-ncov/prepare/transmission.html.

8. Chan JF, et al., A familial cluster of pneumonia associated with the 2019 novel coronavirus indicating person-to-person transmission: a study of a family cluster. Lancet, 2020. 395(10223): p. 514–523.

9. Zhang S, et al., Estimation of the reproductive number of novel coronavirus (COVID- 19) and the probable outbreak size on the Diamond Princess cruise ship: A data-driven analysis. International Journal of Infectious Diseases, 2020. 93: p. 201–204.

10. Rodriguez-Morales AJ, et al., Clinical, laboratory and imaging features of COVID- 19: A systematic review and meta-analysis. Travel Medicine and Infectious Disease, 2020: p. 101623.

11. Liu H, et al., Why are pregnant women susceptible to viral infection: an immunological viewpoint? Journal of Reproductive Immunology, 2020: p. 103122.

12. Wong SF, et al., Pregnancy and perinatal outcomes of women with severe acute respiratory syndrome. Am J Obstet Gynecol, 2004. 191(1): p. 292–7.

13. Wang D, et al., Clinical Characteristics of 138 Hospitalized Patients With 2019 Novel Coronavirus–Infected Pneumonia in Wuhan, China. JAMA, 2020. 323(11): p. 1061–1069.

14. Bastola A, et al., The first 2019 novel coronavirus case in Nepal. Lancet Infect Dis, 2020. 20(3): p. 279–280.

15. Cheng S-C, et al., First case of Coronavirus Disease 2019 (COVID-19) pneumonia in Taiwan. Journal of the Formosan Medical Association, 2020. 119(3): p. 747–751.

16. Holshue ML, et al., First Case of 2019 Novel Coronavirus in the United States. New England Journal of Medicine, 2020. 382(10): p. 929–936.

17. Kim JY, et al., The First Case of 2019 Novel Coronavirus Pneumonia Imported into Korea from Wuhan, China: Implication for Infection Prevention and Control Measures. J Korean Med Sci, 2020. 35(5).

18. Silverstein WK, et al., First imported case of 2019 novel coronavirus in Canada, presenting as mild pneumonia. The Lancet, 2020. 395(10225): p. 734.

19. Liu H, et al., Clinical and CT Imaging Features of the COVID-19 Pneumonia: Focus on Pregnant Women and Children. Journal of Infection, 2020. 11: p. 11.

20. Zhu H, et al., Clinical analysis of 10 neonates born to mothers with 2019-nCoV pneumonia. Translational pediatrics, 2020. 9(1): p. 51–60.

21. Liu D, et al., Pregnancy and Perinatal Outcomes of Women With Coronavirus Disease (COVID-19) Pneumonia: A Preliminary Analysis. AJR, 2020. American Journal of Roentgenology.: p. 1–6.

22. Moher D, et al., Preferred reporting items for systematic reviews and meta-analyses: the PRISMA statement. PLoS Med, 2009. 6(7): p. e1000097.

23. Jiang F, et al., Review of the Clinical Characteristics of Coronavirus Disease 2019 (COVID-19). Journal of General Internal Medicine, 2020.

24. Song F, et al., Emerging 2019 Novel Coronavirus (2019-nCoV) Pneumonia. Radiology, 2020. 295(1): p. 210–217.

25. Lam CM, et al., A case-controlled study comparing clinical course and outcomes of pregnant and non-pregnant women with severe acute respiratory syndrome. BJOG: An International Journal of Obstetrics & Gynaecology, 2004. 111(8): p. 771–4.

26. Gautret P, et al., Hydroxychloroquine and azithromycin as a treatment of COVID-19: results of an open-label non-randomized clinical trial. International Journal of Antimicrobial Agents, 2020: p. 105949.

27. Levy M, et al., Pregnancy outcome following first trimester exposure to chloroquine. Am J Perinatol, 1991. 8(3): p. 174–8.

28. Transfer of drugs and other chemicals into human milk. Pediatrics, 2001. 108(3): p. 776–89.

29. Boelaert JR, et al., Chloroquine accumulates in breast-milk cells: potential impact in the prophylaxis of postnatal mother-to-child transmission of HIV-1. AIDS, 2001. 15(16): p. 2205–7.

30. Zhou F, et al., Clinical course and risk factors for mortality of adult inpatients with COVID-19 in Wuhan, China: a retrospective cohort study. The Lancet, 2020. 395(10229): p. 1054–1062.

31. Rothan HA and Byrareddy SN, The epidemiology and pathogenesis of coronavirus disease (COVID-19) outbreak. J Autoimmun, 2020: p. 102433.

32. Chen N, et al., Epidemiological and clinical characteristics of 99 cases of 2019 novel coronavirus pneumonia in Wuhan, China: a descriptive study. The Lancet, 2020. 395(10223): p. 507–513.

33. Yu WC, Hui DSC, and Chan-Yeung M, Antiviral agents and corticosteroids in the treatment of severe acute respiratory syndrome (SARS). Thorax, 2004. 59(8): p. 643.

34. Bandoli G, et al., A Review of Systemic Corticosteroid Use in Pregnancy and the Risk of Select Pregnancy and Birth Outcomes. Rheumatic diseases clinics of North America, 2017. 43(3): p. 489–502.

35. De Brouwere V, et al., Need for caesarean sections in west Africa. The Lancet, 2002. 359(9310): p. 974–975.

36. Khan MN, et al., Socio-demographic predictors and average annual rates of caesarean section in Bangladesh between 2004 and 2014. PLOS ONE, 2017. 12(5): p. e0177579.

37. Sharma D and Spearman P, The impact of cesarean delivery on transmission of infectious agents to the neonate. Clin Perinatol, 2008. 35(2): p. 407-20, vii-viii.

38. Huang Q-t, et al., Chronic hepatitis B infection and risk of preterm labor: a meta-analysis of observational studies. Journal of Clinical Virology, 2014. 61(1): p. 3–8.

39. Huang Q, et al., The risk of placental abruption and placenta previa in pregnant women with chronic hepatitis B viral infection: a systematic review and meta-analysis. Placenta, 2014. 35(8): p. 539–545.

40. Uddin SMI, et al., Burden and Risk Factors for Coronavirus Infections in Infants in Rural Nepal. Clinical Infectious Diseases, 2018. 67(10): p. 1507–1514.

41. Gagneur A, et al., Coronavirus-related nosocomial viral respiratory infections in a neonatal and paediatric intensive care unit: a prospective study. Journal of Hospital Infection, 2002. 51(1): p. 59–64.

42. Racicot K and Mor G, Risks associated with viral infections during pregnancy. The Journal of clinical investigation, 2017. 127(5): p. 1591–1599.

43. Chen H, et al., Clinical characteristics and intrauterine vertical transmission potential of COVID-19 infection in nine pregnant women: a retrospective review of medical records. Lancet, 2020. 395(10226): p. 809–815.

44. Liu Y, et al., Clinical manifestations and outcome of SARS-CoV-2 infection during pregnancy. Journal of Infection, 2020. 04: p. 04.

45. Wang X, et al., A case of 2019 Novel Coronavirus in a pregnant woman with preterm delivery. Clinical Infectious Diseases, 2020. 28: p. 28.

46. Chen Y, et al., Infants Born to Mothers With a New Coronavirus (COVID-19). Frontiers in Pediatrics, 2020. 8(104).

47. Yu N, et al., Clinical features and obstetric and neonatal outcomes of pregnant patients with COVID-19 in Wuhan, China: a retrospective, single-centre, descriptive study. The Lancet Infectious Diseases.

48. Zambrano LI, et al., A pregnant woman with COVID-19 in Central America. Travel Medicine and Infectious Disease, 2020: p. 101639.

